# Trends in telehealth use by Medicare fee-for-service beneficiaries and its impact on overall volume of healthcare services

**DOI:** 10.1101/2022.06.15.22276468

**Authors:** Chad Ellimoottil, Ziwei Zhu, Rodney L. Dunn, Michael P. Thompson

## Abstract

**Introduction:** At the start of the COVID-19 public health emergency, the federal government made temporary Medicare policy changes to expand telehealth coverage, resulting in a surge in telehealth use. As federal and state policymakers currently consider permanent telehealth policy options, it is important to understand the trends in telehealth use during 2021 and whether telehealth has led to an increase in the overall volume of healthcare services.

**Methods:** Our analysis was conducted using Part B claims for 100% of Medicare fee-for-service beneficiaries. We identified all outpatient evaluation and management (E&M) services received by beneficiaries from January 1, 2019 through December 31, 2021. We then calculated the monthly proportion of outpatient E&M services that were performed in-person and through telehealth.

**Results:** The total number of all outpatient E&M services was 289.0 million in 2019, 255.2 million in 2020 (11.7% lower than 2019), and 260.7 million in 2021 (9.8% lower than 2019). Monthly telehealth services peaked at 7.2 million (or 50.7% of monthly E&M services) in April 2020, followed by a slow decline through the end of 2021. During the second half of 2021, telehealth services made up 8.5-9.5% of monthly E&M services.

**Conclusion:** From April 2020 through December 2021, the monthly volume of telehealth services slowly declined and has plateaued between 8.5-9.5% of all outpatient E&M services received by Medicare fee-for-service beneficiaries. Importantly, the total volume of outpatient E&M services was lower in 2020 and 2021, suggesting that the COVID-19 telehealth flexibilities have not increased the overall volume of outpatient E&M services received by Medicare beneficiaries. These findings should mitigate some concerns about the impact of telehealth on overall healthcare utilization.

At the start of the COVID-19 public health emergency, the federal government made temporary Medicare policy changes to expand telehealth coverage, resulting in a surge in telehealth use.^1,2^ While telehealth was a necessary substitute for in-person care during first few months of the pandemic, there was a decline in the use of telehealth during the second half of 2020.^3^ As federal and state policymakers currently consider permanent telehealth policy options, it is important to understand the trends in telehealth use during 2021 and whether telehealth has led to an increase in the overall volume of healthcare services.

## METHODS

Our analysis was conducted using Part B claims for 100% of Medicare fee-for-service beneficiaries. For each year of our study, we excluded patients who were not enrolled in Medicare Part B or were enrolled in Medicare Advantage. For our first analysis, we identified all outpatient evaluation and management (E&M) services received by beneficiaries from January 1, 2019 through December 31, 2021. We identified telehealth services using Medicare’s list of eligible telehealth services and the appropriate modifier codes (GT, GQ, 95) or place of service code (02) corresponding to each year of the study. We also identified telehealth services using Healthcare Common Procedure Coding System codes for selected virtual care services including phone visits, virtual check-ins, online digital evaluations, interprofessional consultations, and remote monitoring. We then calculated the monthly proportion of outpatient E&M services that were performed in-person and through telehealth.

For our second analysis, we categorized all beneficiaries who received outpatient E&M services from January 1, 2021 through December 31, 2021 as *telehealth users* if they received at least one telehealth service and *telehealth non-users* if they did not receive any telehealth services. We then compared the characteristics of telehealth users and non-users. We used SAS Enterprise Guide (version 7.15) and the institutional review board determined the study was exempt.

## RESULTS

The total number of all outpatient E&M services was 289.0 million in 2019, 255.2 million in 2020 (11.7% lower than 2019), and 260.7 million in 2021 (9.8% lower than 2019). Monthly telehealth services peaked at 7.2 million (or 50.7% of monthly E&M services) in April 2020, followed by a slow decline through the end of 2021 (Figure 1). During the second half of 2021, telehealth services made up 8.5-9.5% of monthly E&M services.

**Figure 1:**
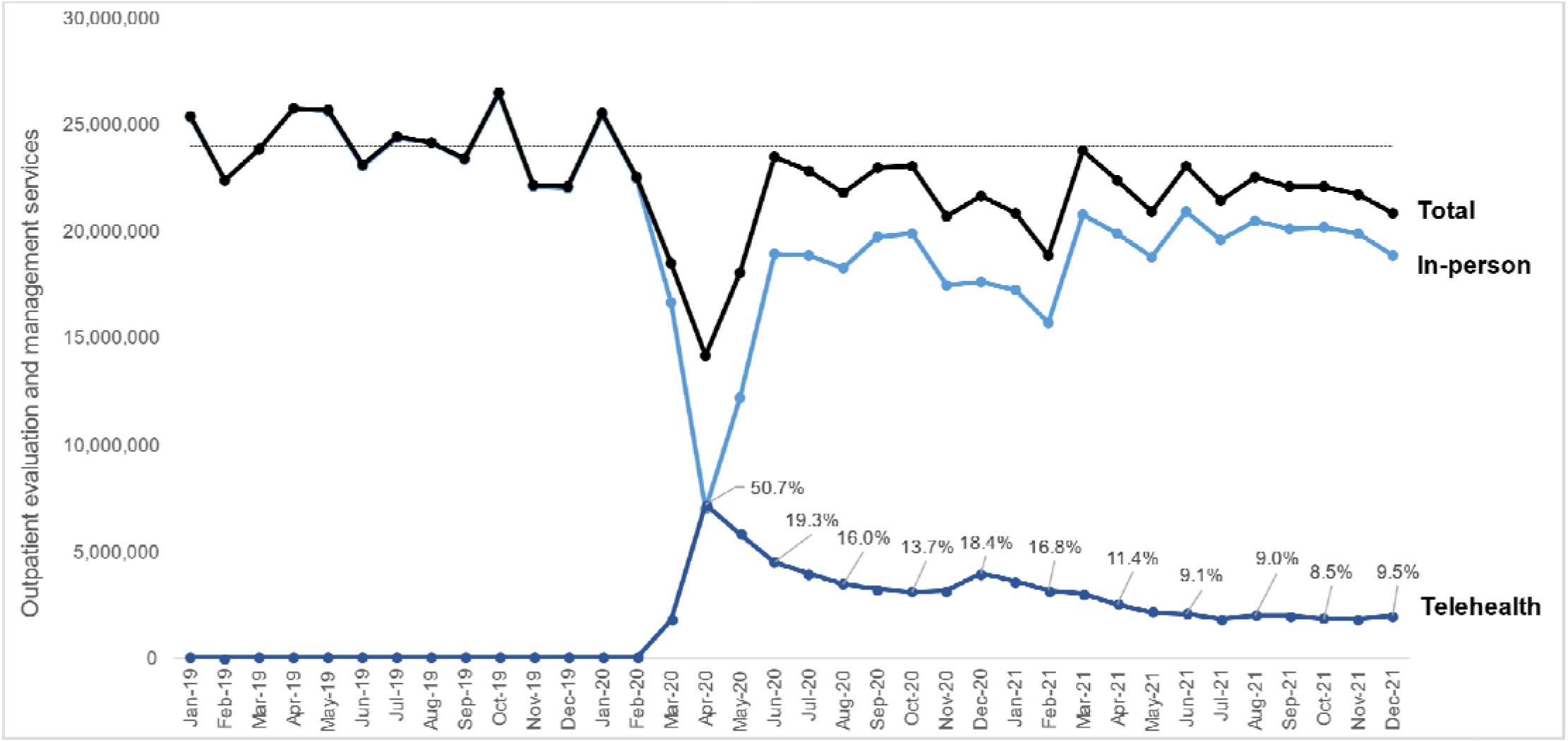
Monthly trend in outpatient evaluation and management (E&M) services for Medicare beneficiaries. Evaluation of 100% of Medicare fee-for-service beneficiaries from January 1, 2019 through December 31, 2021. Telehealth includes outpatient services such as office visits, phone visits, virtual check-in, online digital evaluation, remote monitoring, and interprofessional consult. Median number of monthly E&M services from January 1, 2019 through December 31, 2019 was 24,026,954.

In 2021, 33% of Medicare beneficiaries received at least one telehealth service. (Table 1) Rural beneficiaries utilized telehealth less often than their non-rural counterparts (17% vs 26%). Only minor differences were noted between telehealth users and non-users in regard to age, gender, race and Medicaid eligibility (dual-eligible) status.

**Table 1:** Characteristics of telehealth non-users telehealth users and non-users.

|  |  | No.(%)<sup>a</sup> |  |
| --- | --- | --- | --- |
| Characteristics<sup>b</sup> |  | Telehealth user<sup>c</sup><br>(n=8,711,138) | Telehealth non-user<sup>d</sup><br>(n=17,340,831) |
| Age (%) | <=65 | 1,423,413 (16) | 1,604,561 (9) |
|  | 66-70 | 2,039,508 (23) | 4,555,880 (26) |
|  | 71-75 | 2,009,357 (23) | 4,306,413 (25) |
|  | 76-80 | 1,419,013 (16) | 2,973,040 (17) |
|  | >80 | 1,819,847 (21) | 3,900,937 (22) |
| Sex | Male | 3,571,575 (41) | 7,842,372 (45) |
|  | Female | 5,139,562 (59) | 9,498,456 (55) |
| Race<sup>e</sup> | Non-hispanic white | 6,878,453 (79) | 14,319,833 (83) |
|  | Black (or African-American) | 683,034 (8) | 1,198,704 (7) |
|  | Asian/Pacific islander | 293,663 (3) | 445,782 (3) |
|  | Hispanic | 524,869 (6) | 778,494 (4) |
|  | American Indian/Alaska Native | 50,454 (0.6) | 78,994 (0.5) |
|  | Other/Unknown | 280,665 (3) | 519,024 (3) |
| Zip code | Rural<sup>f</sup> | 1,512,446 (17) | 4,587,707 (26) |
|  | Non-rural | 7,198,692 (83) | 12,753,124 (74) |
| Dual eligibility<sup>g</sup> | Yes | 1,851,119 (21) | 2,388,395 (14) |
|  | No | 6,860,019 (79) | 14,952,436 (86) |

<sup>a</sup>Percentages may not equal 100 due to rounding<sup>b</sup>Analysis of 100% Medicare fee-for-service beneficiaries from January 1, 2021 through December 31, 2021.<sup>c</sup>Telehealth users are defined as beneficiaries who received at least one telehealth visit during 2021. Telehealth includes outpatient services such as office visits, phone visits, virtual check-in, online digital evaluation, remote monitoring, interprofessional consult.<sup>d</sup>Telehealth non-users received no telehealth services during 2021<sup>e</sup>Race was defined using Research Triangle Institute race code in the Master Beneficiary Summary File<sup>f</sup>Rural was defined using Federal Office of Rural Health Policy (FORHP) Data Files<sup>g</sup>Dual eligible beneficiaries were defined as those who had more than one month of eligibility in 2021.

## DISCUSSION

From April 2020 through December 2021, the monthly volume of telehealth services slowly declined and has plateaued between 8.5-9.5% of all outpatient E&M services received by Medicare fee-for-service beneficiaries. Importantly, the total volume of outpatient E&M services was lower in 2020 and 2021, suggesting that the COVID-19 telehealth flexibilities have not increased the overall volume of outpatient E&M services received by Medicare beneficiaries. These findings should mitigate some concerns about the impact of telehealth on overall healthcare utilization. This study was limited by the fact that it did not include a small percent of claims received more than three months after the end of 2021.

## Grant support

1 K08 HS027632-01 (Ellimoottil)

1R01HS028397-01A1 Agency for Healthcare Research and Quality (Ellimoottil and Thompson)

University of Michigan Faculty Catalyst (Ellimoottil)

## Data Availability

All data in the present study are publically available through the Chronic Conditions Data Warehouse

